# Hyperbaric Oxygen for Treatment of Long COVID Syndrome (HOT-LoCO); Protocol for a Randomised, Placebo-Controlled, Double-Blind, Phase II Clinical Trial

**DOI:** 10.1101/2022.05.20.22275312

**Authors:** Anders Kjellberg, Lina Abdel-Halim, Adrian Hassler, Sara El Gharbi, Sarah Al-Ezerjawi, Emil Boström, Carl Johan Sundberg, John Pernow, Koshiar Medson, Jan Kowalski, Kenny A Rodriguez-Wallberg, Xiaowei Zheng, Sergiu-Bogdan Catrina, Michael Runold, Marcus Ståhlberg, Judith Bruchfeld, Malin Nygren-Bonnier, Peter Lindholm

## Abstract

**Introduction:** Long COVID, where symptoms persist 12 weeks after the initial SARS-CoV-2-infection, is a substantial problem for individuals and society in the surge of the pandemic. Common symptoms are fatigue, post-exertional malaise, and cognitive dysfunction. There is currently no effective treatment, and the underlying mechanisms are unknown although several hypotheses exist, with chronic inflammation as a common denominator. In prospective studies, hyperbaric oxygen therapy (HBOT) has been suggested to be effective for the treatment of similar syndromes such as chronic fatigue syndrome and fibromyalgia. A case series has suggested positive effects of HBOT in Long COVID. This randomised placebo-controlled clinical trial will explore HBOT as a potential treatment for Long COVID. The primary objective is to evaluate if HBOT improves health related quality of life (HRQoL) for patients with Long COVID compared to placebo/sham. The main secondary objectives are to evaluate whether HBOT improves endothelial function, objective physical performance, and short term HRQoL.

**Methods and Analysis:** A randomised, placebo-controlled, double-blind, phase II clinical trial in 80 previously healthy subjects debilitated due to Long COVID, with low HRQoL. Clinical data, HRQoL- questionnaires, blood samples, objective tests and activity meter data will be collected at baseline. Subjects will be randomised to a maximum of 10 treatments with hyperbaric oxygen or sham treatment over six weeks. Assessments for safety and efficacy will be performed at six, 13, 26 and 52 weeks, with the primary endpoint (physical domains in RAND-36) and main secondary endpoints defined at 13 weeks after baseline. Data will be reviewed by an independent Data Safety Monitoring Board.

**Ethics and Dissemination:** The trial is approved by The Swedish National Institutional Review Board (2021-02634) and the Swedish Medical Product Agency (5.1-2020-36673). Positive, negative, and inconclusive results will be published in peer-reviewed scientific journals with open access.

**Trial Registration:** NCT04842448. EudraCT: 2021-000764-30

**Strengths and limitations of this trial:** Strengths

- Randomised placebo-controlled, double-blind, parallel groups, clinical trial in compliance with ICH-GCP
- Evaluation of safety and efficacy, including objective and explanatory endpoints
- Independent Data Safety Monitoring Board (DSMB)

Limitations

- New syndrome with unknown mechanisms
- Power calculation is based on similar syndromes
- Selection bias as patients are enrolled from the same post-COVID clinic

## Introduction/Background

In the wake of the first wave of the SARS-CoV-2 pandemic, a new set of often debilitating post-infectious symptoms have arisen. Such symptoms that persist for more than three months, even after mild SARS-CoV infection, have become a major burden for the individuals affected, health care providers, and society in general^1^. The prevalence of long COVID is difficult to determine due to a plethora of symptoms and different definitions^2^. A recent estimation from a UK cohort of 508,707 patients suggests that more than 30% had experienced at least one symptom with “significant impact on my daily life” giving an overall prevalence of 1.72%^3^. Most patients experiencing lingering symptoms are women, of which many have experienced only mild if any respiratory symptoms, and seldom required hospital care during the acute phase of their SARS-CoV-2 infection ^4^. Reported long-term symptoms include shortness of breath, fatigue, post-exertional malaise, and cognitive dysfunction, frequently leading to reduced working capability ^2^. Some patients are also diagnosed with autonomic dysfunction, including Postural Orthostatic Tachycardia Syndrome (POTS) and inappropriate sinus tachycardia^5 6^.

As the pandemic continues to spread, with new mutations and resulting variants of SARS-CoV-2 appearing, effective treatments are needed to quell infection rates as well as mitigate lingering long-term symptoms. There is still not a uniform definition or name of the syndrome, but post-acute COVID-19 syndrome (PACS), post COVID syndrome (PCS), or Long COVID are commonly used^7^. An attempt to achieve a global definition of Post COVID condition, the name suggested by World Health organisation (WHO), was recently made by a Delphi consensus process^8^. Post COVID condition is previously listed in International Classification of Diseases (ICD-10) with code U09.9, which includes all commonly used names. Experts in the field have recently suggested management guidelines for monitoring and follow-up, but to date there is no effective treatment^9^. The underlying mechanisms are not understood but several hypotheses including endothelial dysfunction, oxidative stress, and chronic inflammation have been proposed^10 11^. In fact, a recent study demonstrated persistent microvascular endothelial dysfunction for four months following COVID-19 infection^12^.

Hyperbaric oxygen therapy (HBOT) is administered by delivering 100% oxygen at raised pressure to patients in a hyperbaric chamber. HBOT has previously been used as an adjunctive treatment for COVID-19, resulting in faster recovery in prospective trials, case series^13^, and a randomised controlled trial (RCT)^14^, with additional RCTs ongoing^15^. The rationale for treatment of COVID-19 with HBOT is the treatment’s well-established anti-inflammatory effects^16 17^. Furthermore, a small retrospective cohort study has shown promising results in alleviating symptoms of Long COVID in patients treated with HBOT^18^. The safety profile of HBOT is well established and is considered both safe and effective for the treatment of several chronic inflammatory diseases such as soft tissue radiation injury^19^. HBOT has been shown to improve symptoms and quality of life in other syndromes associated with chronic fatigue^20 21^. We explore HBOT administered within a randomised placebo-controlled clinical trial as a potential treatment for patients suffering from Long COVID. The purpose of this manuscript is to provide a summary of our protocol that complies with International Council for Harmonisation-Good Clinical Practice (ICH-GCP), with a detailed description and rationale for the primary and main secondary endpoints, including patient reported outcomes (PRO) in line with Standard Protocol Items: Recommendations for Interventional Trials (SPIRIT) SPIRIT-PRO Extension Guidelines^22^.

### Hypothesis and objectives

The overall hypothesis to be evaluated is that HBOT reduces oxidative stress and chronic inflammation, improves endothelial dysfunction, and thereby alleviates symptoms associated with Long COVID.

The primary objective is to evaluate whether HBOT improves Health related quality of life (HRQoL) for patients compared to placebo. The main secondary objectives are to evaluate whether HBOT improves endothelial dysfunction, objective physical performance, and improvement of short term HRQoL. Other secondary objectives are to evaluate if HBOT improves autonomic dysfunction, restorative sleep, the health-economic benefits of the treatment and evaluate biomarkers for the HBO effect on inflammation and chronic hypoxia. Furthermore, we aim to evaluate the safety profile of HBOT for Long COVID patients.

## Methods and analysis

### Trial design

The trial is designed as a prospective, randomised, placebo-controlled, double-blind, phase II clinical trial. The trial consists of 5 visits for 52 weeks. At Visit 1 the participant eligibility will be established, and baseline data collected. Block randomisation will be performed, stratified by gender and disease severity as determined by the RAND-36-questionnaire. Eligible subjects are randomised a maximum of two weeks before the first treatment and will receive a maximum of ten treatments over six weeks from randomisation. Treatment is conducted by designated staff not involved in assessment or data collection, subjects and investigators are blinded to the treatment allocation. The randomisation and blinding process is described in a standard operating procedure (SOP). Visit 2 is conducted on the day of the last treatment. The primary and main secondary endpoints will be assessed at 13 weeks from baseline at Visit 3. Visits 4 and 5 are long term follow-up. Subjects will also be asked to participate in a post-trial follow up over 4 years. A flowchart of the trial design is depicted in Figure 1. and the Consolidated Standards of Trials (CONSORT) flow diagram is depicted in Figure 2.

**Figure 1.**
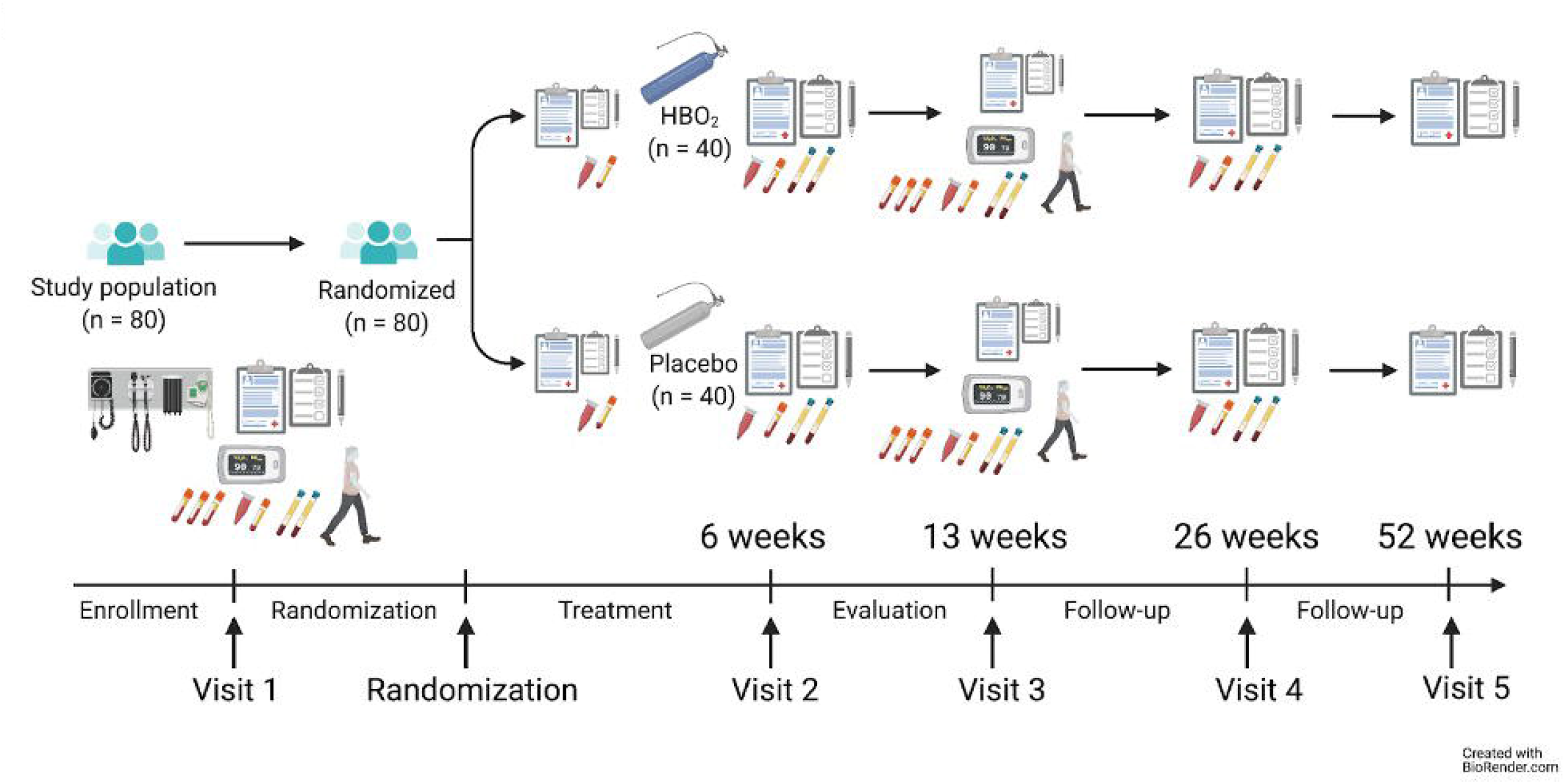
Trial Flowchart.

**Figure 2.**
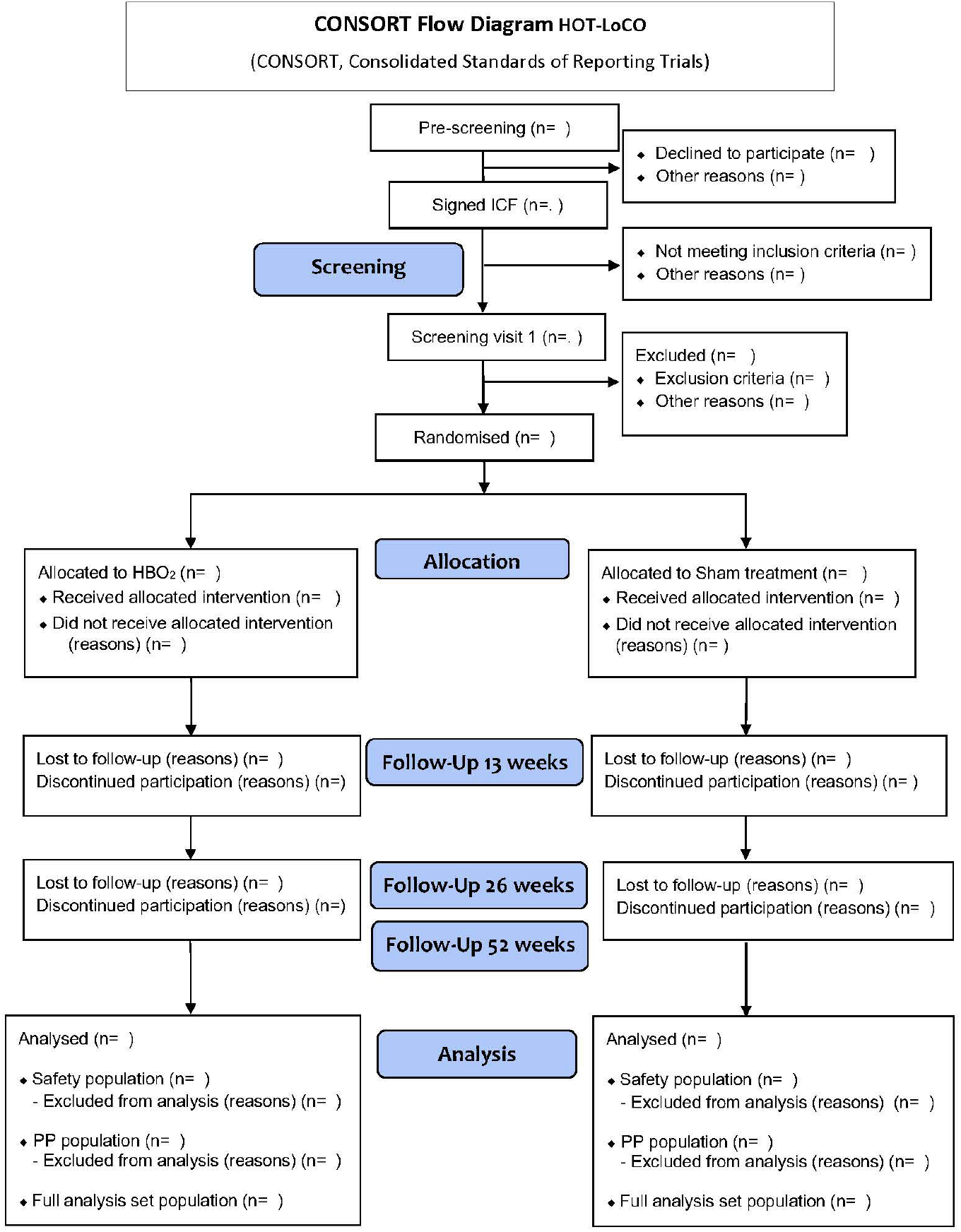
Consolidated Standards of Trials (CONSORT) flow diagram.

#### Patient and Public Involvement

The trial design and consent form were discussed with and approved by a patient representative. We thank Svenska Covidföreningen through chairman Åsa Kristofferson-Hedlund for their support.

### Setting

The trial is investigator initiated and will take place in a single center. The sponsor is Region Stockholm via the Karolinska University Hospital in collaboration with Karolinska Institutet, both in Stockholm, Sweden. Patients will be recruited through the post-COVID outpatient clinic and/or advertisement. Measurements and treatments will take place at the hyperbaric unit. If included in the trial, all patients regardless of intervention or control will be treated at the hyperbaric treatment facility, staffed by anesthesiologists and intensivists as well as nurses specifically trained in HBOT. All personnel involved in the trial are designated to specific duties and trained in ICH-GCP. As per protocol at Karolinska University Hospital, each treatment in the hyperbaric chambers must be overseen by a minimum of two staff members. Local, national, and international guidelines for clinical trials and HBOT during the COVID-19 pandemic will be followed.

### Trial population

80 patients aged 18–60, previously generally healthy (defined as American Society of Anesthesiologists (ASA) class I-II), will be recruited. They must have had symptoms consistent with Long COVID for a minimum of 12 weeks, as well as a Long COVID diagnosis with ICD- 10 code U09.9. Subjects must have been working or studying before the diagnosis. A HBOT specific questionnaire with focus on HBOT contraindications will be filled in by all subjects, contraindications include pregnancy, claustrophobia, obstructive lung disease and history of spontaneous pneumothorax. All inclusion and exclusion criteria are listed in Table 1. Subjects who are diagnosed with Long COVID through the Karolinska University Hospital Post-COVID outpatient clinic will be evaluated by a multidisciplinary team consisting of an infectious disease specialist, pulmonologist, cardiologist as well as a physiotherapist. All patients will be assessed with a battery of questionnaires, physical tests, laboratory tests and radiology including MRI’s.

**Table 1.** Inclusion and exclusion criteria.

| Inclusion criteria | Exclusion criteria |
| --- | --- |
| • Aged 18–60 years<br>• Healthy or mild systemic disease (ASA I-II) prior to COVID-19<br>• Symptoms consistent with Long COVID for at least 12 weeks<br>• Diagnosed with Long COVID, PACS, PCS (ICD-10 U09.9)<br>• Working or studying prior to COVID-19<br>• Documented informed consent according to ICH-GCP and national regulations | • Known pregnancy or positive pregnancy test in women of childbearing age<br>• ASA III or more from other cause than Long COVID<br>• Score above 70 in RAND-36 Role Limitation Physical Health (RP) or Physical Functioning (PF)<br>• Diabetes mellitus<br>• Diagnosed with hypertension prior to COVID-19<br>• Contraindication for HBOT treatment according to local guidelines<br>• Participation or recent participation in a clinical trial with an investigational product<br>• Mental inability, reluctance or language |
|  | difficulties that result in difficulty understanding the meaning of trial participation |

### Treatment/interventions

The HBOT group will undergo HBOT at 2.4 Atmospheres absolute (ATA), approximately 240kPa for 90 minutes with two airbrakes, with a maximum of 10 treatments within 6 weeks of randomisation. The placebo group will undergo ‘Sham treatment’ with air-breathing at 1.34 ATA, approximately 134kPa to equate the sensation of HBOT and airbrakes will be simulated. They will undergo a maximum of 10 treatments within 6 weeks of randomisation. Both treatment protocols and Blinding SOP are available as supplementary material.

The hyperbaric chambers to be used are designed for a single patient (monoplace chamber) or for multiple patients (multi-place chamber). In the case of the monoplace chamber, it is pressurized with 100% oxygen and staff and equipment are located outside the chamber. However, multi-place chambers are pressurized with air, allowing staff and equipment to be inside the same chamber where the patient breathes oxygen through a mask. The latter is suitable for patients requiring a high level of medical care or groups of patients that can sit in a chair for 90 minutes, whereas the monoplace chamber is more comfortable but requires the patient to be fully alert and stable.

### Procedures

The patients will be informed about the trial orally and in writing and given the chance to ask questions. If they agree to participate, an informed consent form (ICF) will be signed by the patient and an investigator before any study-specific procedures occur. Subjects will then be scheduled for a screening visit (Visit 1) where baseline data will be collected, and inclusion/exclusion criteria are verified. Subjects eligible for inclusion in the trial will subsequently enter the trial, be randomised, and allocated to treatment. After the treatment period of six weeks, the subjects will be scheduled for follow-up visits at 13 +/- 2 weeks and 26 and 52 weeks +/- 4 weeks after randomisation.

All procedures in the trial are described in detail in the full protocol that is availible as supplementary material. For treatments, blinding procedures, and assessments, standard operating procedures (SOPs) will be followed. A list of procedures is depicted in Table 2.

**Table 2.** List of procedures. (Trial specific procedures are marked with **bold X**, data collected from medical records are marked with narrow X) *Exclusion criteria includes a pregnancy test (if applicable), RAND 36 questionnaire, a HBOT specific questionnaire, review of medical records and a medical examination if needed. ** Medical history includes COVID-19 specific history, routine blood tests, questionnaires, physical tests, and radiology, medical records will be reviewed and recorded. *** Socio-demography that may change over time such as sick-leave, weight, activity, smoking habits.

| Activity | Visit 1 | Visit 2 | Visit 3 | Visit 4 | Visit 5 |
| --- | --- | --- | --- | --- | --- |
| Week | 0 | 6 | 13 | 26 | 52 |
| **Signed Informed consent Form** | **X** |  |  |  |  |
| **Inclusion/exclusion criteria** | **X\*** |  |  |  |  |
| **Randomisation** | **X** |  |  |  |  |
| **Medical history** | **X** | **X\*\*** | **X\*\*** | **X\*\*** | **X\*\*** |
| **Socio-demography** | **X** | **X\*\*\*** | **X\*\*\*** | **X\*\*\*** | **X\*\*\*** |
| **Concomitant medications** | **X** | **X** | **X** | **X** | **X** |
| **RAND 36** | X | X | X | X | X |
| **EQ-5D** | X | X | X | X | X |
| **RHI** | X |  | X |  |  |
| **6 min walk test** | X | X | X | X | X |
| **30/60 s chair-stand** | X | X | X | X |  |
| **Nexfin** | X |  | X |  |  |
| **Treatment (HBOT/Placebo)** |  | X (1-10) |  |  |  |
| **Treatment planned** |  | X (1-10) |  |  |  |
| **AE/ADR** | X | X | X | X | X |
| **Trial-specific biochemistry** | X | X | X | X | X |
| **Biobanking (PBMC, Plasma, EPR)** | X | X, X | X | X |  |
| **Activity meter** | X | X | X | X | X |

### Assessments/measurements

Prior to inclusion subjects will have undergone extensive tests, including radiology with different modalities such as computer tomography (CT), magnetic resonance imaging (MRI), dual-energy computer tomography (DECT), cardiac ultrasound and chest X-rays, and objective physical measurements such as handgrip strength, spirometry and head-up-tilt test and questionnaires used in clinical practice to confirm the diagnosis and rule out any differential diagnosis. This data will be obtained from medical records.

Blood-based biochemical values for kidney function, liver function, cardiac enzymes, haematology, and blood glucose will be obtained from patients’ medical records. Trial-specific biochemistry will include ferritin, D-dimer, LDH, troponin T, and a pregnancy test for any woman of childbearing age; blood for biobanking will be collected from fasting subjects.

During the screening visit (Visit 1) subjects will fill out the RAND 36-item Health Survey (RAND-36), EuroQol-5 Dimensions Questionnaire (EQ-5D) and undergo physical tests including the 6-minute walk test (6MWT) and 30/60 sec chair stand test (CST), and other objective evaluations including endothelial function with pulse amplitude tonometry (PAT), measurements of cardiac function, and activity, heart rate variability and sleep patterns with an activity meter.

### Patient Reported Outcome (PRO) Measures

#### RAND-36-item health survey (RAND-36)

RAND 36 is a self-reporting questionnaire that contains 36 items that measure eight concepts of health in general terms, at present and past four weeks: physical functioning (ten items), role limitations due to physical health (four items), role limitations due to emotional problems (three items), energy/fatigue (four items), emotional well-being (five items), social functioning (two items), pain (two items) and general health (five items). It also includes a single item that provides an indication of perceived change in health over the last year. Scoring RAND 36 is a two-step process. First, numeric values from the survey are coded so that all items are scored from 0 (lowest score) to 100 (highest possible score). Scores then represent the percentage of total possible score achieved. In step two, items in the same scale are averaged together to create the eight-scale scores. Items that are left blank (missing data) are not considered when calculating the scale scores. Hence, scale scores represent the average for all items in the scale that the respondent answered. RAND 36 is well documented in terms of reliability and variability also for Swedish translation ^23^. National gender and age normative data are availible for comparison^23^ The questionnaire will be sent out digitally to the subjects on the day of the visit, and when filled out uploaded to the medical records. The dimensions in RAND-36 are presented separately and we have chosen the physical domains RP and PF as primary endpoint for two reasons:

1. The physical domains seem to be severely affected in conditions associated with chronic fatigue and POTS^24 25^.
2. We expect the physical domains to be least affected by placebo.

#### EuroQol-5 Dimensions Questionnaire (EQ-5D)

EQ-5D is a widely used patient-reported questionnaire aimed at measuring five different dimensions of present health with three or five levels of severity: no problems, some/moderate problems, and severe/extreme problems. The five different dimensions are mobility, self-care, usual activities, pain/discomfort, anxiety/depression. It also uses a visual analogue scale (VAS) 0-100 for quantifying measures of overall health. EQ-5D is a well-validated tool and the index that is calculated from the dimensions gives an estimate of Quality Adjusted Life Years (QALY), with a low index indicating a low HRQoL^26^. We will use five levels of severity (EQ-5D-5L) in our trial. One of the strengths of EQ-5D is that gender and age normative data for the Swedish population is available for use in health economic evaluation^27^, and the index can be used to predict ability to work or study. The questionnaire will be sent out digitally to the subjects on the day of the visit and when filled out, uploaded to the medical records.

The rationale for choosing RAND-36 is that it is well validated and used in previous studies with similar methodology to enable power calculations. EQ-5D was chosen to provide an evaluation of HRQoL in a shorter perspective, as it is easier to fill in and may therefore be a better option for long term follow-up, to enable a simple health economic evaluation.

### Physical tests

#### 6-minute walk test (6MWT)

The 6MWT will be performed in a corridor with a measured distance of 30 m, with markings for every meter. The subject will carry a pulse oximeter with a probe attached to their forehead. The test will be monitored by an experienced instructor recording parameters every minute, the total number of meters walked in six minutes, the subject’s graded and subjective feeling of leg-fatigue and dyspnea according to the Borg CR-10-scale, as well as the feeling of general exertion according to the Borg-RPE-scale, both at baseline and at the end of the tests^28^.

#### 30/60 seconds chair stand test (CST)

Here the subject will stand up straight and sit down completely as many times as possible for 30/60 seconds (s). An instructor will record the number of times the subject manages to perform the movement, as well as the subject’s graded and subjective feeling of general exertion according to the Borg-RPE-Scale, and dyspnea and leg fatigue according to the Borg CR-10-scale at baseline and the end of the test. The rationale for recording 30/60 s is that some subjects may not be able to perform the full 60 s test.

### Objective measurements

#### Nexfin

The Nexfin monitor will be connected to a fasting subject. This is a non-invasive measurement of cardiovascular indices, with a beat-to-beat pulse wave analyzer. The Nexfin device (ClearSight, Edwards Lifesciences) is placed on the middle phalanx of the middle finger on the right hand. The Nexfin device comprises a pneumatic plethysmograph that provides advanced hemodynamic parameters and continuous noninvasive blood pressure (BP) from a finger cuff, with a redesigned self-coiling mechanism that reconstructs the clinical standard brachial arterial waveform from the finger arterial pressure waveform; it has been validated towards invasive measurements in several clinical trials^29^.

#### Reactive Hyperemia Index (RHI)

Endothelial function will be determined in fasting state using an EndoPAT 2000 device (Itamar Medical, Caesarea, Israel). The subjects will be connected to the pulse amplitude tonometry (PAT) device for non-invasive determination of digital endothelial function. The PAT device comprises a pneumatic plethysmograph that allows measurements of pulse amplitude at baseline and during hyperemia following a five minutes arterial occlusion of the forearm ^30^. The change in the PAT signal is used for calculating the reactive hyperemia index (RHI), which has been shown to reflect microvascular endothelial dysfunction, reduced NO bioavailability and to predict cardiovascular events ^31^.

#### Activity meter

The OURA™ ring (Oura Health Oy) will be used as an activity tracker that registers heart rate variability, body temperature, physical activity, and sleep patterns. Subjects will wear the ring for at least 1 week before and after each visit with data being synced in OURA’s smartphone application which subsequently will be uploaded to an encrypted database ^32^. The weekly mean of each variable will be collected.

### Randomisation

Subjects who meet the inclusion criteria will be randomised using a digital tool, Randomizer.at, version 2.0.0 (Institute for Medical Informatics, Statistics and Documentation, Medical University of Graz). The system has a complete electronic audit trail for all activities involved with the randomisation. Randomisation is stratified for gender and ‘illness severity’. Illness severity is determined as the mean of RAND-36 score for RP and PF into three strata: 1. <30, 2. 30-50 and 3. >50. Investigators access the randomisation system through a web portal with access control. Staff designated to treatment allocation have user-specific access to the unblinded treatment schedule. Study treatment is allocated according to protocol, 10 treatments over six weeks, a maximum of two weeks after randomisation.

Subjects as well as all personnel participating in assessments of symptoms and any objective findings will be blinded to the treatment. The placebo ‘Sham treatment’ protocol is well established and even experienced divers cannot differ between Sham treatment and HBOT ^33^. Designated personnel, experienced in HBOT and trained in GCP and the specific protocols will administer the assigned treatments. All subjects will furthermore be asked during the first week of treatment whether they believe they received the placebo treatment or HBOT, to validate the blinding process.

### Trial endpoints

The primary endpoints are the mean change from baseline to 13 weeks in RAND 36 domains RP and PF respectively. The main secondary endpoints are mean change from baseline to 13 weeks in RHI, 6MWT, 30/60 s CST, EQ-5D and proportion of subjects with a normalisation of levels in RAND-36 domains RP and PF respectively, at 13 weeks. Primary-, Main secondary-, Selected other- and Safety endpoints are listed in **Table 3**.

**Table 3.** HOT-LoCO: Trial endpoints.

|  |  |
| --- | --- |
| Primary endpoints | Mean change from baseline to 13 weeks in RAND 36 domains role limitations due to physical health and physical functioning respectively. |
| Main secondary efficacy endpoints | I. Mean change from baseline to 13 weeks in RHI.<br>II. Mean change from baseline to 13 weeks in the 6MWT.<br>III. Mean change from baseline to 13 weeks in the |
|  | 30/60 s CST.<br>IV. Mean change from baseline to 13 weeks in EQ-5D scores.<br>V. Proportion of subjects with a normalisation* of levels in RAND-36 domains RP and PF respectively, at 13 weeks. |
| Other efficacy endpoints | I. Mean change in other RAND-36 domains at 13, 26 and 52 weeks compared to baseline.<br>II. Mean change in EQ-5D at 6, 26 and 52 weeks compared to baseline.<br>III. Mean change in physical activity using an activity meter at 6, 13 and 26 weeks compared to baseline<br>IV. Mean change in HRV using an activity meter at 6, 13 and 26 weeks compared to baseline<br>V. Mean change in sleeping pattern using an activity meter at 6, 13 and 26 weeks compared to baseline. |
| Explorative/Descriptive endpoints | I. Mean change from baseline in hypoxia pathways in PBMCs evaluated by RNA sequencing, at 6, 13 and 26 weeks.<br>II. Mean change from baseline in inflammatory PBMCs evaluated by RNA sequencing, at 6, 13 and 26 weeks<br>III. Mean change from baseline of reactive oxygen species in red blood cells measured by EPR, at, 6 and 13 weeks.<br>IV. Mean change from baseline of microRNA in plasma, at 6 and 13 weeks.<br>V. Mean change from baseline in trial-specific clinical biochemistry at 6 and 13 weeks.<br>a. D-Dimer<br>b. Ferritin<br>c. LDH<br>d. Troponin T<br>VI. Long term post-trial follow-up of HRQoL using EQ-5D as variable up to 4 years post trial. |
| Safety and compliance endpoints | I. Number of subjects, proportion of subjects and number of adverse events (AEs) at 13 weeks.<br>II. Number of subjects, proportion of subjects that have completed planned treatments after 6 weeks. |

\* According to Swedish normative data<sup>23</sup>

### Safety and adverse events

Collection of Adverse events (AE) and Serious Adverse Events (SAE) data will start directly after inclusion and will be recorded until Visit 3. Only SAE will be collected outside the treatment period (Visit 2). Ongoing AE and SAE at the end of Visit 3 will be followed up during long-term follow-up until the subject’s last visit. The definition, handling, follow-up, and reporting of AEs are defined in the original protocol (p.34–38). The safety endpoints will be evaluated by an independent Data Safety Monitoring Board (DSMB) in the context of the trial design and currently existing information about Long COVID and HBOT. The DSMB is composed of three experts in their respective disciplines of medicine, clinical trial methodology and conduct. The DSMB will review the data at the predetermined interim analyses and at the end of trial, a charter delineating their guidelines for operating and stopping rules for terminating individual subjects, a portion or all the trial prematurely, was drawn up and agreed upon before the trial started. The members of the DSMB, meeting plan and responsibilities are specified in the original protocol (p.6 and 44).

### Statistical analyses

This section is a short summary of the planned statistical analyses of the most important endpoints including primary and main secondary endpoints. A longer summary is availible in the full protocol (p.38-42). A more technical and detailed elaboration of the principal features will be written in a separate Statistical Analysis Plan (SAP). The SAP will be finalised prior to Data Base Lock (DBL).

### Analysis population

Full analysis set (FAS), per-protocol (PP) and safety population (SP) will be employed. The FAS population will be defined as: all randomized subjects who were exposed at least once to the study intervention.

### Sample size calculation

The primary endpoint is mean change from baseline to week 13 in the RAND-36 score. A ten-point higher mean change in the HBOT group compared to the placebo group is considered as a clinically relevant difference. Sample size calculation was performed using t-test for independent groups, with an 80% power), and a type-I error rate of 0.05 (5%), assuming a common SD of 15 from prior studies, to detect a 10-unit difference between groups. Power calculations indicates that at least 37 subjects per group are needed. Subsequently, we aim to recruit 80 subjects. nQuery, version 7 was used for sample size calculation.

### Hypothesis testing and adjustment for multiplicity

Hypothesis testing will be controlled at the type-I error rate of 0.05 and adequately adjusted for multiplicity in the two primary endpoints. However, there will be no adjustment for multiplicity in main secondary endpoints as this is an exploratory study, but nominal p-values will be presented, and results will be interpreted as exploratory findings. All hypothesis tests will be two-sided. Details of the multiplicity adjustment in terms of the selection of endpoints to include in the testing sequence and the criteria for rejecting (or not rejecting) individual hypotheses will be provided in the SAP.

### Subgroups

Subgroup analysis will be done and presented for gender and disease severity defined as the mean of RAND-36 RP and PF and divided into ‘RP and PF below 30’, ‘RP and PF 30-50’ and ‘RP and PF above 50’.

### Statistical methodology

Primary and secondary endpoints will be evaluated using the FAS population and sensitivity analyses performed using the PP population. The primary objective of the study is to confirm a superior efficacy for the active treatment compared to placebo in the primary endpoints. The null hypothesis to be tested is that there is no difference between HBO treatment and placebo, i.e., the mean change in (HBOT) = mean change (placebo). The same statistical hypothesis will be used for key secondary endpoints.

All continuous variables will be described using standard statistical measures, i.e., number of observations, mean and median value, standard deviation, minimum and maximum value. All categorical variables will be summarised in frequency tables.

In general, for continuous outcome variables including the primary endpoint, they will be analysed using ANCOVA, unless otherwise specified, including stratification factors and treatment as fixed factors in the model. Estimates will be presented using least-square means for differences between treatment arms. In addition, continuous endpoints measured repeatedly over time, such as EQ5D and RAND-36 domains, the change from baseline will be analyzed using a linear mixed-effect model including baseline, treatment group, sex, symptom severity, visit, and treatment group by visit interaction, and subjects as random effects, in the models. An unstructured covariance matrix will be assumed.

Analysis for categorical data in terms of binary data (Yes/No) will be presented as the proportion of subjects with the frequency of presence or absence, by treatment group of the characteristics of interest and analysed using the CMH Chi-square test including stratification factors, where the parameter used for the statistical hypothesis testing will be the odds ratio (OR), as a measure of the relative difference in odds between treatment arms. An OR>1 indicates efficacy in favor of HBOT compared to placebo.

Missing data will be adequately imputed for all subjects in the FAS population. In addition, the observed cases population will be evaluated as a sensitivity analysis.

An interim safety analysis will be performed when 20 subjects have available data for the safety endpoints, and a second interim analysis when 40 subjects have data available for primary endpoint to adjust the sample size if needed. The trial will also be evaluated for futility regarding the primary endpoints, i.e., the predictive probability of success at the end of the trial.

### Safety analysis

The number and percentage of patients reporting AEs, and the number of AEs reported will be presented. The events will be tabulated by system organ class and preferred term by treatment group. In addition, summaries by relationship to trial drug and severity will be presented. AEs will also be presented in separate tabulations.

The number of patients experiencing an AE will be compared descriptively between groups. All patients with AEs will be listed individually with the patient number in addition to the type of event, start and stop time, duration, seriousness, severity, any action taken, relationship to trial drug and outcome of AE.

## Discussion

This manuscript presents the trial design and rationale for the HOT-LOCO trial. The trial is conducted in compliance with ICH-GCP to protect the safety and well-being of the subjects as well as the integrity and validity of the data. HBOT has been used for almost a century for other chronic inflammatory conditions with well documented safety profiles for accepted indications ^34^. However, the intervention is not without risk. The nature of the disease, which provokes multiple symptoms and a low quality of life make the risk group a ‘vulnerable group’ and it is important to make sure that the subjects are not unduly influenced by the expectation or benefits associated with participation.

The randomised, double-blinded design is gold standard, and thus is a strength considering primary endpoints being PRO. The trial design involves multiple exploratory and descriptive endpoints, which may provide valuable data regarding the disease regardless of clinical outcomes. Should HBOT prove clinically effective for the efficacy endpoints the trial design also allows further investigation into possible causal mechanisms.

### Limitations

The current trial has some important limitations. Long COVID is a novel disease with unknown mechanisms. The prevalence is continuously being revised and it is not known how symptoms and best practice treatment will evolve over time. The treatment protocol in this trial is novel and thus considered a limitation. Normally, HBOT is administered five days a week, with 30–40 sessions over six to eight weeks. The protocol in this trial is based on experience from severe COVID-19 where five treatments seem to be sufficient. However, more research on the dose is needed. Further limitations lie in the possible selection bias of patients being referred through the same outpatient clinic; most patients are severely debilitated (a prerequisite for referral was at least 50% sick-leave) and due to long waiting times, most patients have been ill for more than one year. The power calculation for the primary endpoint is extrapolated from studies of similar design and diseases with similar symptoms but have not been based on a pilot trial and thus is considered as an increased risk of type II error. However, interim analyses will be performed when 20 patients have data available for safety endpoints, and when 40 patients have available for primary endpoint to minimize the risk of an underpowered trial. Furthermore, ‘sham treatment’ may have up to 58% efficacy^35^. We did not take this into account when we performed our power calculation, which could result in the trial being underpowered. Both EQ-5D and RAND-36 are the most widely used PRO measures for HRQoL and have been used in the setting of long COVID and similar conditions such as ME/CFS and fibromyalgia but due to the novelty of the condition we do not know what to expect from our population and our ‘clinically relevant’ estimation may be set too high. Three to five points have been proposed as a minimally clinically important difference (MCID) for RAND-36 when used in health economic evaluation^36^. This assumption in our power calculation may also cause a type II error.

### Ethics and dissemination

The trial is conducted in accordance with The Declaration of Helsinki, ICH-GCP, local and national regulations. The trial was approved by The Swedish ethical review board (EPM no 2021-02634, amendment 2021-04572), approval 2021-05-25 and 2021-09-22 and The Swedish medical products agency (LV no 5.1-2020-36673), approval 2021-07-06. The trial was registered online (NCT04842448) and EudraCT number: 2021-000764-30 before start of the trial.

The trial is monitored by the Karolinska Trial Alliance (KTA) before the trial started, during the trial, and after trial completion. A designated monitor will monitor the randomisation and blinding process. The monitoring is performed to ensure that the trial is conducted in compliance with the protocol, detailed in a separate monitoring plan and that data is handled according to ICH-GCP.

The first publication will report the results of the interim safety analysis to help other researchers in trial designs and health care providers in decision making. The main publication will report the primary and main secondary endpoints together with the full safety and compliance report at 13 weeks. Separate publications will report exploratory endpoints: 1. Descriptive results from the Ouraring, 2. Health economic analysis, 3. Exploratory biomarkers and biochemical analyses. 4. Descriptive results from medical history that is collected during the trial 5. Depending on the outcome of the primary endpoint at 13 weeks, follow-up on HRQoL at 26 and 52 weeks. 6. Long time, post-trial follow-up on HRQoL, 4 years.

### Current trial status

The first subject was included in September 2021. Nineteen subjects have been randomized, 14 have completed the intervention by February 1, 2022. The first safety analysis will be performed when 20 subjects have completed the interventions, according to the plan Q1 2022.

## Supporting information

SPIRIT Checklist

## Data Availability

The full trial protocol, statistical plan and consent form will be publicly available. Data will be available on patient level; data will be pseudonymised, the full dataset and statistical code will be available upon request. All publications will be made available on Open Access. Source data will be described in a Meta-data repository. A full description of the intended use of the data must be sent to the corresponding author for review and approval. Participant consent for data sharing is conditioned and new ethics approval may be required.

## Authors contribution

AK is the principal investigator who wrote the hypothesis and developed most of the protocol together with PL. AK and PL wrote the applications to Swedish IRB and MPA. LAH drafted the manuscript together with AK. AH, SEG, SAE and EB are sub-investigators, enrolling and evaluating subjects and collecting data. MNB, JB, MS, and MR are trial chairs involved in writing the protocol and applications. JK wrote the statistical analysis plan together with AK and designed the randomisation. All authors including CJS, KRW, SBC, XZ and JP contributed to the current submission and critically reviewed the manuscript. AK is corresponding author for this work and attests that all listed authors meet authorship criteria and that no others meeting the criteria have been omitted.

### Funding

This project is funded by The Swedish Heart-Lung foundation (HLF), Stockholm Health Council (ALF) and Oura Health Oy. The funding bodies are not involved in the study design, collection of data, or in the forthcoming analyses and interpretation of data, writing of manuscripts or in the decisions to submit manuscripts for publication.

### Patient consent for publication

Not required.

## Acknowledgements

Study coordinator Felicia Doeser for invaluable help with managing subjects and collecting data. The doctors and nurses at the hyperbaric unit at Karolinska University Hospital involved in the treatments and allocation to the treatment groups; Doctors: Karl-Fredrik Sjölund, Johan Thelaus and Georgios Sidiras Nurses: Carola Lernbäck, Birgitta Johansson and Johan Ohlberger and Annelie Kruthammar. Medical student: Lovisa Liwenborg. The director of Intensive care, Björn Persson, director of Health professions, Emma Sjölund and director of Cardiology Frieder Braunschweig for supporting this project. The research nurses at KFE for planning and help with blood sampling; Anna Schening, Anna Granström, Ola Friman and Pia Zetterqvist. Physiotherapists Anna Svensson-Raskh and Ulrika Holdar for planning and performing the physical tests. Staff at Studiecenter Karolinska for setting up the laboratory manual and handling blood samples.

## References

1. Goertz YMJ, Van Herck M, Delbressine JM, et al. Persistent symptoms 3 months after a SARS-CoV-2 infection: the post-COVID-19 syndrome? ERJ Open Res 2020;6(4) doi: 10.1183/23120541.00542-2020 [published Online First: 2020/12/02]

2. Deer RR, Rock MA, Vasilevsky N, et al. Characterizing Long COVID: Deep Phenotype of a Complex Condition. EBioMedicine 2021;74:103722. doi: 10.1016/j.ebiom.2021.103722 [published Online First: 2021/11/29]

3. Whitaker M. Persistent symptoms following SARS-CoV-2 infection in a random community sample of 508,707 people 2021 [Available from: https://spiral.imperial.ac.uk/handle/10044/1/89844 accessed 9-Jan-2022 2022.

4. Davis HE, Assaf GS, McCorkell L, et al. Characterizing long COVID in an international cohort: 7 months of symptoms and their impact. EClinicalMedicine 2021;38:101019. doi: 10.1016/j.eclinm.2021.101019 [published Online First: 2021/07/27]

5. Johansson M, Stahlberg M, Runold M, et al. Long-Haul Post-COVID-19 Symptoms Presenting as a Variant of Postural Orthostatic Tachycardia Syndrome: The Swedish Experience. JACC Case Rep 2021;3(4):573–80. doi: 10.1016/j.jaccas.2021.01.009 [published Online First: 2021/03/17]

6. Stahlberg M, Reistam U, Fedorowski A, et al. Post-Covid-19 Tachycardia Syndrome: A distinct phenotype of Post-acute Covid-19 Syndrome. Am J Med 2021 doi: 10.1016/j.amjmed.2021.07.004 [published Online First: 2021/08/15]

7. Venkatesan P. NICE guideline on long COVID. The lancet Respiratory medicine 2021 doi: 10.1016/S2213-2600(21)00031-X [published Online First: 2021/01/17]

8. Soriano JB, Murthy S, Marshall JC, et al. A clinical case definition of post-COVID-19 condition by a Delphi consensus. Lancet Infect Dis 2021 doi: 10.1016/S1473-3099(21)00703-9 [published Online First: 2021/12/25]

9. Shah W, Hillman T, Playford ED, et al. Managing the long term effects of covid-19: summary of NICE, SIGN, and RCGP rapid guideline. Bmj 2021;372:136. doi: 10.1136/bmj.n136 [published Online First: 2021/01/24]

10. Ferraro E, Germano M, Mollace R, et al. HIF-1, the Warburg Effect, and Macrophage/Microglia Polarization Potential Role in COVID-19 Pathogenesis. Oxid Med Cell Longev 2021;2021:8841911. doi: 10.1155/2021/8841911 [published Online First: 2021/04/06]

11. Chang R, Mamun A, Dominic A, et al. SARS-CoV-2 Mediated Endothelial Dysfunction: The Potential Role of Chronic Oxidative Stress. Frontiers in physiology 2020;11:605908. doi: 10.3389/fphys.2020.605908 [published Online First: 2021/02/02]

12. Mahdi A, Collado A, Tengbom J, et al. Erythrocytes Induce Vascular Dysfunction in COVID-19. In: Institutet K, ed. Preprint ed. JACC: Basic to Translational Science: SSRN, 2021.

13. Oliaei S, SeyedAlinaghi S, Mehrtak M, et al. The effects of hyperbaric oxygen therapy (HBOT) on coronavirus disease-2019 (COVID-19): a systematic review. Eur J Med Res 2021;26(1):96. doi: 10.1186/s40001-021-00570-2 [published Online First: 2021/08/21]

14. Cannellotto M, Duarte M, Keller G, et al. Hyperbaric oxygen as an adjuvant treatment for patients with COVID-19 severe hypoxaemia: a randomised controlled trial. Emergency medicine journal : EMJ 2021 doi: 10.1136/emermed-2021-211253 [published Online First: 2021/12/16]

15. Kjellberg A, Douglas J, Pawlik MT, et al. Randomised, controlled, open label, multicentre clinical trial to explore safety and efficacy of hyperbaric oxygen for preventing ICU admission, morbidity and mortality in adult patients with COVID-19. BMJ Open 2021;11(7):e046738. doi: 10.1136/bmjopen-2020-046738 [published Online First: 2021/07/07]

16. Kjellberg A, De Maio A, Lindholm P. Can hyperbaric oxygen safely serve as an anti-inflammatory treatment for COVID-19? Medical Hypotheses 2020;144 doi: 10.1016/j.mehy.2020.110224 [published Online First: 30 Aug]

17. De Maio A, Hightower LE. COVID-19, acute respiratory distress syndrome (ARDS), and hyperbaric oxygen therapy (HBOT): what is the link? Cell Stress Chaperones 2020:1–4. doi: 10.1007/s12192-020-01121-0 [published Online First: 2020/05/20]

18. Robbins T, Gonevski M, Clark C, et al. Hyperbaric oxygen therapy for the treatment of long COVID: early evaluation of a highly promising intervention. Clin Med (Lond) 2021;21(6):e629–e32. doi: 10.7861/clinmed.2021-0462 [published Online First: 2021/12/05]

19. Oscarsson N, Muller B, Rosen A, et al. Radiation-induced cystitis treated with hyperbaric oxygen therapy (RICH-ART): a randomised, controlled, phase 2-3 trial. The lancet oncology 2019;20(11):1602–14. doi: 10.1016/S1470-2045(19)30494-2 [published Online First: 2019/09/21]

20. Efrati S, Golan H, Bechor Y, et al. Hyperbaric oxygen therapy can diminish fibromyalgia syndrome--prospective clinical trial. PloS one 2015;10(5):e0127012. doi: 10.1371/journal.pone.0127012 [published Online First: 2015/05/27]

21. Akarsu S, Tekin L, Ay H, et al. The efficacy of hyperbaric oxygen therapy in the management of chronic fatigue syndrome. Undersea Hyperb Med 2013;40(2):197–200. [published Online First: 2013/05/21]

22. Calvert M, Kyte D, Mercieca-Bebber R, et al. Guidelines for Inclusion of Patient-Reported Outcomes in Clinical Trial Protocols: The SPIRIT-PRO Extension. JAMA : the journal of the American Medical Association 2018;319(5):483–94. doi: 10.1001/jama.2017.21903 [published Online First: 2018/02/08]

23. Orwelius L, Nilsson M, Nilsson E, et al. The Swedish RAND-36 Health Survey - reliability and responsiveness assessed in patient populations using Svensson’s method for paired ordinal data. J Patient Rep Outcomes 2017;2(1):4. doi: 10.1186/s41687-018-0030-0 [published Online First: 2017/01/01]

24. Hardt J, Buchwald D, Wilks D, et al. Health-related quality of life in patients with chronic fatigue syndrome: an international study. J Psychosom Res 2001;51(2):431–4. doi: 10.1016/s0022-3999(01)00220-3 [published Online First: 2001/08/23]

25. Bagai K, Song Y, Ling JF, et al. Sleep disturbances and diminished quality of life in postural tachycardia syndrome. J Clin Sleep Med 2011;7(2):204–10. [published Online First: 2011/04/22]

26. Dolan P. Modeling valuations for EuroQol health states. Med Care 1997;35(11):1095–108. doi: 10.1097/00005650-199711000-00002 [published Online First: 1997/11/21]

27. Burstrom K, Sun S, Gerdtham UG, et al. Swedish experience-based value sets for EQ-5D health states. Qual Life Res 2014;23(2):431–42. doi: 10.1007/s11136-013-0496-4 [published Online First: 2013/08/27]

28. Enright PL. The six-minute walk test. Respiratory care 2003;48(8):783–5. [published Online First: 2003/08/02]

29. Ameloot K, Van De Vijver K, Broch O, et al. Nexfin noninvasive continuous hemodynamic monitoring: validation against continuous pulse contour and intermittent transpulmonary thermodilution derived cardiac output in critically ill patients. ScientificWorldJournal 2013;2013:519080. doi: 10.1155/2013/519080 [published Online First: 2013/12/10]

30. Hamburg NM, Benjamin EJ. Assessment of endothelial function using digital pulse amplitude tonometry. Trends Cardiovasc Med 2009;19(1):6–11. doi: 10.1016/j.tcm.2009.03.001 [published Online First: 2009/05/27]

31. Alexander Y, Osto E, Schmidt-Trucksass A, et al. Endothelial function in cardiovascular medicine: a consensus paper of the European Society of Cardiology Working Groups on Atherosclerosis and Vascular Biology, Aorta and Peripheral Vascular Diseases, Coronary Pathophysiology and Microcirculation, and Thrombosis. Cardiovasc Res 2021;117(1):29–42. doi: 10.1093/cvr/cvaa085 [published Online First: 2020/04/14]

32. Altini M, Kinnunen H. The Promise of Sleep: A Multi-Sensor Approach for Accurate Sleep Stage Detection Using the Oura Ring. Sensors (Basel) 2021;21(13) doi: 10.3390/s21134302 [published Online First: 2021/07/03]

33. Lansdorp CA, van Hulst RA. Double-blind trials in hyperbaric medicine: A narrative review on past experiences and considerations in designing sham hyperbaric treatment. Clin Trials 2018;15(5):462–76. doi: 10.1177/1740774518776952 [published Online First: 2018/06/06]

34. Heyboer M, 3rd, Sharma D, Santiago W, et al. Hyperbaric Oxygen Therapy: Side Effects Defined and Quantified. Adv Wound Care (New Rochelle) 2017;6(6):210–24. doi: 10.1089/wound.2016.0718 [published Online First: 2017/06/16]

35. Redberg RF. Sham controls in medical device trials. The New England journal of medicine 2014;371(10):892–3. doi: 10.1056/NEJMp1406388 [published Online First: 2014/09/04]

36. Samsa G, Edelman D, Rothman ML, et al. Determining clinically important differences in health status measures: a general approach with illustration to the Health Utilities Index Mark II. Pharmacoeconomics 1999;15(2):141–55. doi: 10.2165/00019053-199915020-00003 [published Online First: 1999/06/03]

